# Heart disease mortality in cancer survivors: A population-based study in Japan

**DOI:** 10.1101/2023.02.23.23286382

**Authors:** Yasufumi Gon, Ling Zha, Tsutomu Sasaki, Toshitaka Morishima, Yuko Ohno, Hideki Mochizuki, Tomotaka Sobue, Isao Miyashiro

**Affiliations:** Department of Neurology, Osaka University Graduate School of Medicine, Suita, Osaka, Japan; Cancer Control Center, Osaka International Cancer Institute, Osaka-shi, Osaka, Japan; Department of Social Medicine, Environmental Medicine and Population Science, Osaka University Graduate School of Medicine, Suita, Osaka, Japan; Department of Mathematical Health Science, Osaka University Graduate School of Medicine, Suita, Osaka, Japan

**Keywords:** cancer, cancer survivors, heart disease, mortality, cohort study

## Abstract

**Background:** The association between cancer survivors and heart disease mortality remains unclear. This study analyzed the risk of fatal heart disease in cancer survivors.

**Methods:** Data from the Osaka Cancer Registry collected between 1985 and 2013 and data from the vital statistics in Japan were retrospectively analyzed. Causes of death were investigated.

Standardized mortality ratios (SMRs) were calculated to compare the risk of fatal heart disease between patients with cancer and the general population. Poisson regression models were used to estimate the risk of fatal heart disease in patients with cancer and other cancer subgroups.

**Results:** In total, 688,473 cancer patients were included in the analysis, and 337,117 patients died during the study period. Among them, 10,781 patients died of heart disease, with 5,020 of these patients having ischemic heart disease; 3,602 patients, heart failure; 441 patients, hypertensive disease; and 1,718 patients, other heart diseases. The SMR (95% confidence interval [CI]) for heart disease was 2.85 (2.80–2.91). The SMRs (95% CIs) for ischemic heart disease, heart failure, and hypertensive disease were 3.26 (3.17–3.35); 2.69 (2.60–2.78), and 4.47 (4.07–4.90), respectively. The risk of fatal heart disease increased over time after cancer diagnosis. The multivariable Poisson regression model showed that males were more likely to die of heart disease than females (relative risk, 1.39; 95% CI, 1.34–1.45). Notably, the risk of fatal heart disease among cancer survivors has decreased in recent years.

**Conclusions:** Cancer survivors have a higher risk of fatal heart disease than the general population.

## Introduction

Advances in cancer care have prolonged survival^1^. With population aging, patients with cancer have also become older and more highly comorbid^2,3^. The number of elderly cancer survivors is expected to increase in the coming decades, and thus, the management of comorbidities is becoming a more significant concern.

Patients with cancer are at a high risk of developing heart disease^4^. Cancer and heart disease share common risk factors such as hypertension, diabetes, and smoking^4,5^. Additionally, neuroendocrine factors, oxidative stress, inflammatory cytokines, and impaired immune system have been implicated in the increased risk of heart disease in patients with cancer ^4,5^. With respect to mortality, an increased risk of death from heart disease has been reported for breast^6,7^, head and neck^8^, testicular^9^, and hematopoietic malignancies^10^. Zaorsky et al. recently found that heart disease was the most common non-cancer cause of death in patients with cancer^11^.

Stoltzfus et al. evaluated the data of approximately 7.5 million cancer patients from the Surveillance, Epidemiology, and End Results (SEER) database and reported a 2.2-fold higher risk of death from heart disease in patients with cancer than in the general population^12^. However, different findings have been reported in literature. A study using the Korean Cancer Registry reported a lower risk of heart disease-related mortality among cancer survivors^13^. Another study using the Tasmanian Cancer Registry demonstrated that the risk of heart disease mortality in patients with cancer was similar to that in the general population^14^.

In an era of increasing numbers of older cancer survivors, clinicians including both oncologists and cardiologists must recognize the significance of the risk of heart disease in patients with cancer. Evidence about the association between cancer and heart disease worldwide needs to be accumulated. However, many of the previous studies have been conducted in Europe and the United States, and only few studies were from Asia^7–14^. Given that Japan has one of the most aged populations worldwide and that one of two Japanese people is predicted to develop cancer^15^, data on the association between cancer and heart disease mortality will be valuable in advancing research in the field of cardio-oncology.

However, the risk of heart disease-related mortality in patients with cancer remains unclear to date. Thus, this study aimed to evaluate the risk of fatal heart disease after cancer diagnosis. We analyzed the risk of fatal heart disease in patients with cancer compared with the general population and other patients with cancer.

## Material and Methods

### Study design and data source

This retrospective study was conducted as part of the Neoplasms ANd other causes of DEath (NANDE) study, which investigated the causes of death in patients with cancer^16-18^. Briefly, the NANDE database was created by linking the Osaka Cancer Registry (OCR) with official statistics in Japan. The OCR has been in operation since 1962, covering ≥ 8,000,000 residents in Osaka Prefecture, Japan. All patients registered in the OCR are followed for 10 years. The OCR includes data on age at diagnosis, sex, year of diagnosis, cancer type, stage at diagnosis, histology, follow-up period, and death. The OCR contains also information on survival or death status, and official statistics in Japan include the individual causes of death based on the death certificate completed by a doctor. Therefore, we merged the two databases by sex, date of birth, date of death, and municipality of residence data and collected details on the causes of death from official statistics. Thereafter, information of 96.6% of the patients with cancer was collated. The NANDE database contains cancer-related information on age at diagnosis, sex, years of diagnosis, cancer type, stage at diagnosis, histology, survival time, and cause of death.

### Patients

The current study used data of patients diagnosed with cancer between 1985 and 2013 and registered in the NANDE database. This cohort was identical to the population previously analyzed for fatal stroke^18^. The exclusion criteria were as follows: (1) uncertain date of death, (2) uncertain final date of survival confirmation, (3) uncertain date of first cancer diagnosis, (4) uncertain date of second cancer diagnosis, (5) uncertain age at first cancer diagnosis, (6) death certificate notification or death certification only, and (7) simultaneous cancer (i.e., multiple tumors identified at the time of diagnosis) or synchronous cancer (i.e., diagnosed within 2 months of each other). The definition of synchronous tumors varies among studies, ranging from 2 to 6 months between diagnoses^19,20^.

### Variable definition

Age at diagnosis was grouped into six categories: ≤39, 40–49, 50–59, 60–69, 70–79, and ≥80 years. Meanwhile, year of diagnosis was classified into three periods: 1985–1995, 1995– 2004, and 2005–2013. The stage at diagnosis was classified into seven categories: (i) intraepithelial (abnormal cells were present but have not spread to nearby tissues), (ii) localized (cancer was limited to the organ where it originated, with no sign of spread), (iii) lymph node metastasis (cancer had spread to regional lymph nodes), (iv) infiltration to adjacent organs (cancer had spread to nearby tissues or organs), (v) distant metastasis (cancer had metastasized to distant parts of the body), (vi) unknown (there is insufficient information to determine the stage), and (vii) not available (missing data on the stage). Histology was classified into five categories according to Berg’s classification^21^: squamous or basal cell carcinoma, adenocarcinoma, other carcinoma, hematopoietic tumor, and other histology. The detailed histological groupings are provided in **Supplementary Table S1**.

Diseases were coded based on the International Classification of Diseases (ICD), and cancers were coded according to the International Classification of Diseases, Oncology, and Oncology Third Edition. Details concerning the assignment of codes and number of patients (those included, excluded, and lost to follow-up) are shown in **Supplementary Table S2**. Heart diseases, including ischemic heart disease, heart failure, hypertensive disease, and other heart diseases, were identified using the ICD-9 and ICD-10 codes. In Japan, the causes of death were officially registered based on the ICD-9 codes from 1979 to 1994; thereafter, the ICD-10 codes were used. Details on the assignment of ICD-9 and ICD-10 codes were as follows: heart disease, ICD-9 (390–429) and ICD-10 (I00–I52); ischemic heart disease, ICD-9 (410–414) and ICD-10 (I20–I25); heart failure, ICD-9 (428) and ICD-10 (I50); hypertensive disease, ICD-9 (401–405) and ICD-10 (I10–I15); and other heart diseases, ICD-9 (390–398, 415–429) and ICD-10 (I00– I09, I26–I49, I51–I52).

### Statistical analysis

The risk of death from heart disease after cancer diagnosis was analyzed. Heart disease death was defined as heart disease being the cause of death recorded on the death certificate. The observation period was from January 1985 to December 2013. The survival time was measured in days, with a minimum of 1 day and a maximum of 3,652 days. Patients diagnosed with a second cancer during the observation period were censored at the time of the second cancer diagnosis.

To compare the risk of fatal heart disease in patients with cancer to that in the general population, the standardized mortality ratios (SMRs) and their 95% confidence intervals (CIs) were calculated as the ratio of the observed to the expected number of deaths. The expected number of deaths was computed using annual Japanese sex- and age-specific death rates. These death rates were calculated by dividing the number of deaths by the national population within each 5-year age group and sex category for a calendar year. Information concerning both the national population and the number of deaths, including in patients with and without cancer, is available on the Portal Site of Official Statistics of Japan (https://www.e-stat.go.jp/en). The SMRs were calculated for heart disease, ischemic heart disease, heart failure, and hypertensive disease.

To compare the risk of fatal heart disease between patients with cancer and various cancer subgroups, Poisson regression models were used to estimate the relative risk (RR) and 95% CIs. The RRs were adjusted for sex, age at diagnosis, period of diagnosis, stage at diagnosis, and histology.

All statistical analyses were performed using Stata 17/MP (StataCorp, College Station, TX, USA) and R (https://cran.r-project.org/) software. All tests were two tailed, and P<0.05 was considered statistically significant.

## Results

### Patient characteristics

Among the 1,007,199 cancer patients initially evaluated, 688,473 patients were included in the analysis, with 2,668,126 person-years at risk. The median (interquartile ranges) follow-up duration was 4.12 (0.99–6.54) years after cancer diagnosis. During the study period, 337,117 patients died; among them, 10,781 patients died of heart disease as follows: ischemic heart disease, 5,020 patients; heart failure, 3,602 patients; hypertensive disease, 441 patients; and other heart diseases, 1,718 patients. The number of deaths due to ischemic heart disease, heart failure, hypertensive disease, and other heart diseases according to cancer type is shown in **Supplemental Table S3**.

### Comparison of risk of heart disease death between cancer patients and the general population

Among all patients with cancer, the crude rate of mortality due to heart disease was 404.07 per 100,000-person-years, and the overall SMR for heart disease was 2.85 (95% CI, 2.80–2.91). **Table 1** presents the SMRs for heart disease death in patients with cancer in comparison to those in the general population. Patients who were diagnosed with heart disease at a younger age had a higher SMR for heart disease, and the SMR gradually decreased as the age of diagnosis increased. The SMR was the highest when the diagnostic period was later than 2005. The SMR by stage at diagnosis was highest for intraepithelial disease and lowest for distant metastatic disease in patients with known stages. SMR by histology was higher in all histologies than in the general population.

**Table 1.**
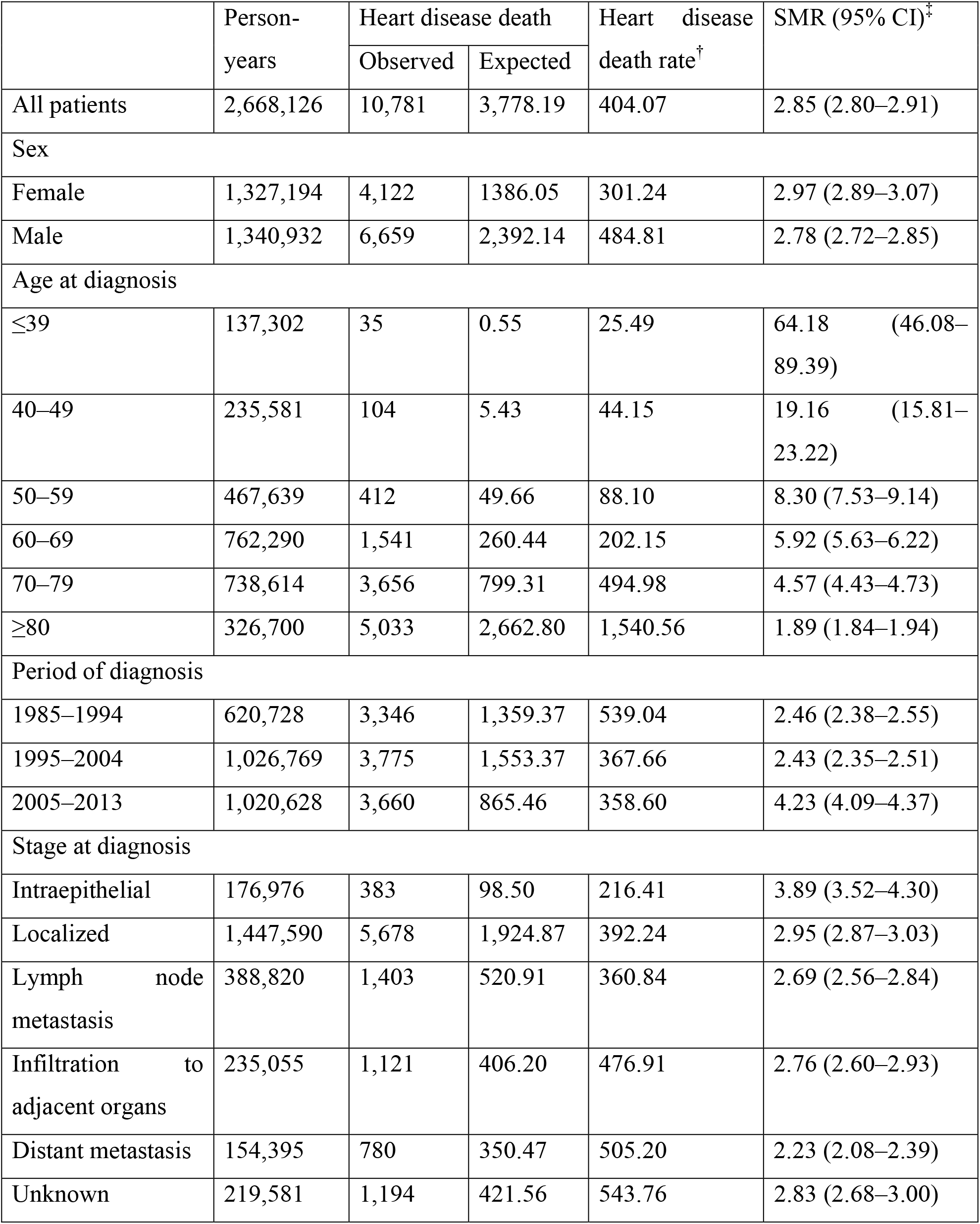
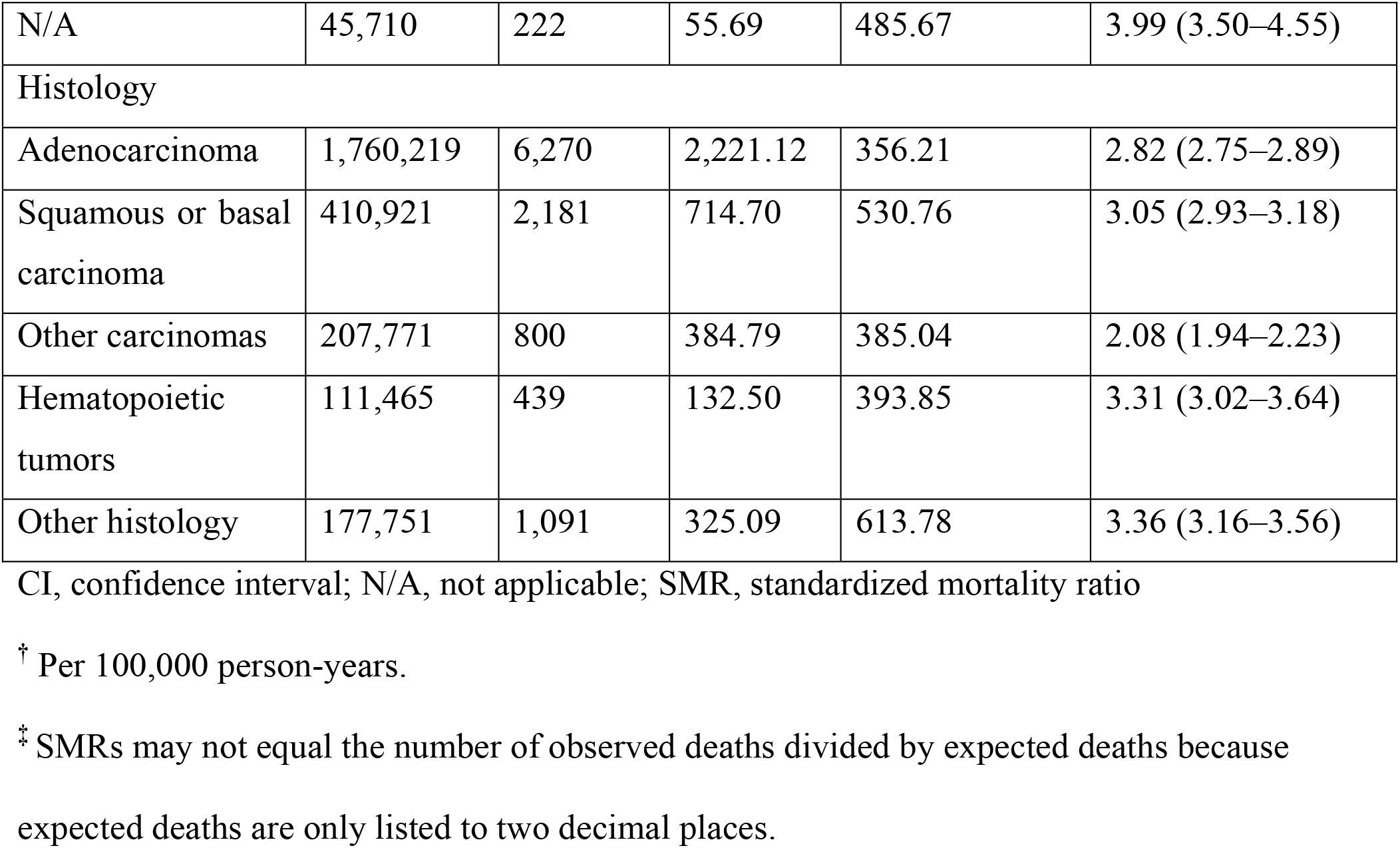
Standardized mortality ratios for heart disease death in patients with cancer compared to the general population.

|  | Person-years | Heart disease death |  | Heart disease death rate<sup>†</sup> | SMR (95% CI)<sup>‡</sup> |
| --- | --- | --- | --- | --- | --- |
|  |  | Observed | Expected |  |  |
| All patients | 2,668,126 | 10,781 | 3,778.19 | 404.07 | 2.85 (2.80–2.91) |
| Sex |  |  |  |  |  |
| Female | 1,327,194 | 4,122 | 1386.05 | 301.24 | 2.97 (2.89–3.07) |
| Male | 1,340,932 | 6,659 | 2,392.14 | 484.81 | 2.78 (2.72–2.85) |
| Age at diagnosis |  |  |  |  |  |
| ≤39 | 137,302 | 35 | 0.55 | 25.49 | 64.18 (46.08–89.39) |
| 40–49 | 235,581 | 104 | 5.43 | 44.15 | 19.16 (15.81–23.22) |
| 50–59 | 467,639 | 412 | 49.66 | 88.10 | 8.30 (7.53–9.14) |
| 60–69 | 762,290 | 1,541 | 260.44 | 202.15 | 5.92 (5.63–6.22) |
| 70–79 | 738,614 | 3,656 | 799.31 | 494.98 | 4.57 (4.43–4.73) |
| ≥80 | 326,700 | 5,033 | 2,662.80 | 1,540.56 | 1.89 (1.84–1.94) |
| Period of diagnosis |  |  |  |  |  |
| 1985–1994 | 620,728 | 3,346 | 1,359.37 | 539.04 | 2.46 (2.38–2.55) |
| 1995–2004 | 1,026,769 | 3,775 | 1,553.37 | 367.66 | 2.43 (2.35–2.51) |
| 2005–2013 | 1,020,628 | 3,660 | 865.46 | 358.60 | 4.23 (4.09–4.37) |
| Stage at diagnosis |  |  |  |  |  |
| Intraepithelial | 176,976 | 383 | 98.50 | 216.41 | 3.89 (3.52–4.30) |
| Localized | 1,447,590 | 5,678 | 1,924.87 | 392.24 | 2.95 (2.87–3.03) |
| Lymph node metastasis | 388,820 | 1,403 | 520.91 | 360.84 | 2.69 (2.56–2.84) |
| Infiltration to adjacent organs | 235,055 | 1,121 | 406.20 | 476.91 | 2.76 (2.60–2.93) |
| Distant metastasis | 154,395 | 780 | 350.47 | 505.20 | 2.23 (2.08–2.39) |
| Unknown | 219,581 | 1,194 | 421.56 | 543.76 | 2.83 (2.68–3.00) |
| N/A | 45,710 | 222 | 55.69 | 485.67 | 3.99 (3.50–4.55) |
| Histology |  |  |  |  |  |
| Adenocarcinoma | 1,760,219 | 6,270 | 2,221.12 | 356.21 | 2.82 (2.75–2.89) |
| Squamous or basal carcinoma | 410,921 | 2,181 | 714.70 | 530.76 | 3.05 (2.93–3.18) |
| Other carcinomas | 207,771 | 800 | 384.79 | 385.04 | 2.08 (1.94–2.23) |
| Hematopoietic tumors | 111,465 | 439 | 132.50 | 393.85 | 3.31 (3.02–3.64) |
| Other histology | 177,751 | 1,091 | 325.09 | 613.78 | 3.36 (3.16–3.56) |

CI, confidence interval; N/A, not applicable; SMR, standardized mortality ratio
<sup>†</sup> Per 100,000 person-years.
<sup>‡</sup> SMRs may not equal the number of observed deaths divided by expected deaths because expected deaths are only listed to two decimal places.

### Comparison of risk of ischemic heart disease, heart failure, and hypertensive disease death between cancer patients and the general population

**Table 2** describes the SMRs for ischemic heart disease, heart failure, and hypertensive disease. The number of deaths due to ischemic heart disease, heart failure, and hypertensive disease is listed in **Supplemental Table S4**. The crude mortality rates due to ischemic heart disease, heart failure, and hypertensive disease were 188.15, 135.00, and 16.53 per 100,000-person-years, respectively. The SMRs for ischemic heart disease, heart failure, and hypertensive disease were 3.26 (95% CI, 3.17–3.35), 2.69 (95% CI, 2.60–2.78), and 4.47 (95% CI, 4.07–4.90), respectively. The SMRs for ischemic heart disease, heart failure, and hypertensive disease were higher in females, those diagnosed between 2005 and 2013, and those with intraepithelial diseases than in other subgroups. Regarding age at diagnosis, the SMRs for ischemic heart disease and heart failure were the highest in patients with a younger age at diagnosis. In contrast, the SMR for hypertensive disease was the highest in patients aged 60–69 years at diagnosis. The SMRs varied among the histological types.

**Table 2.**
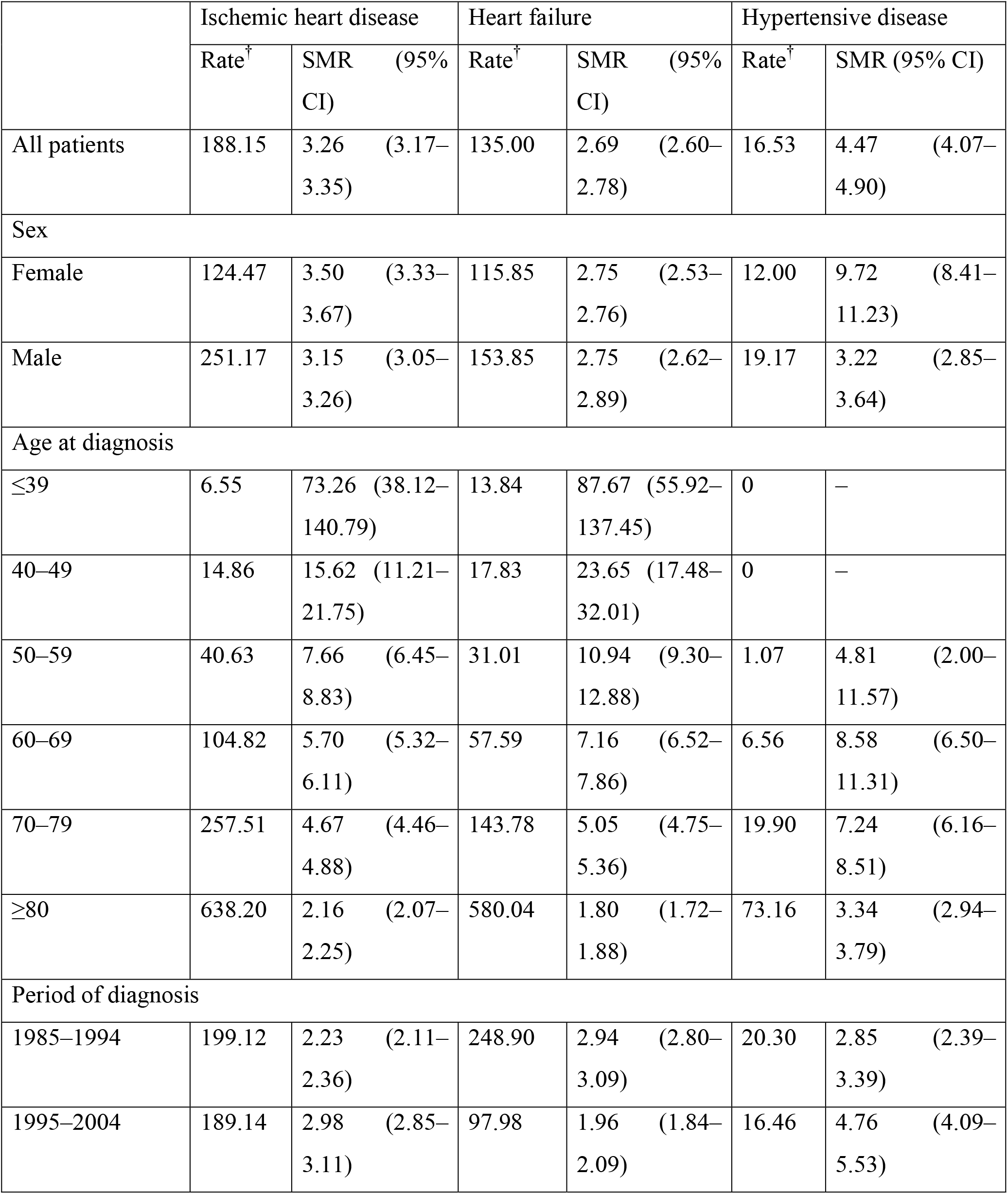
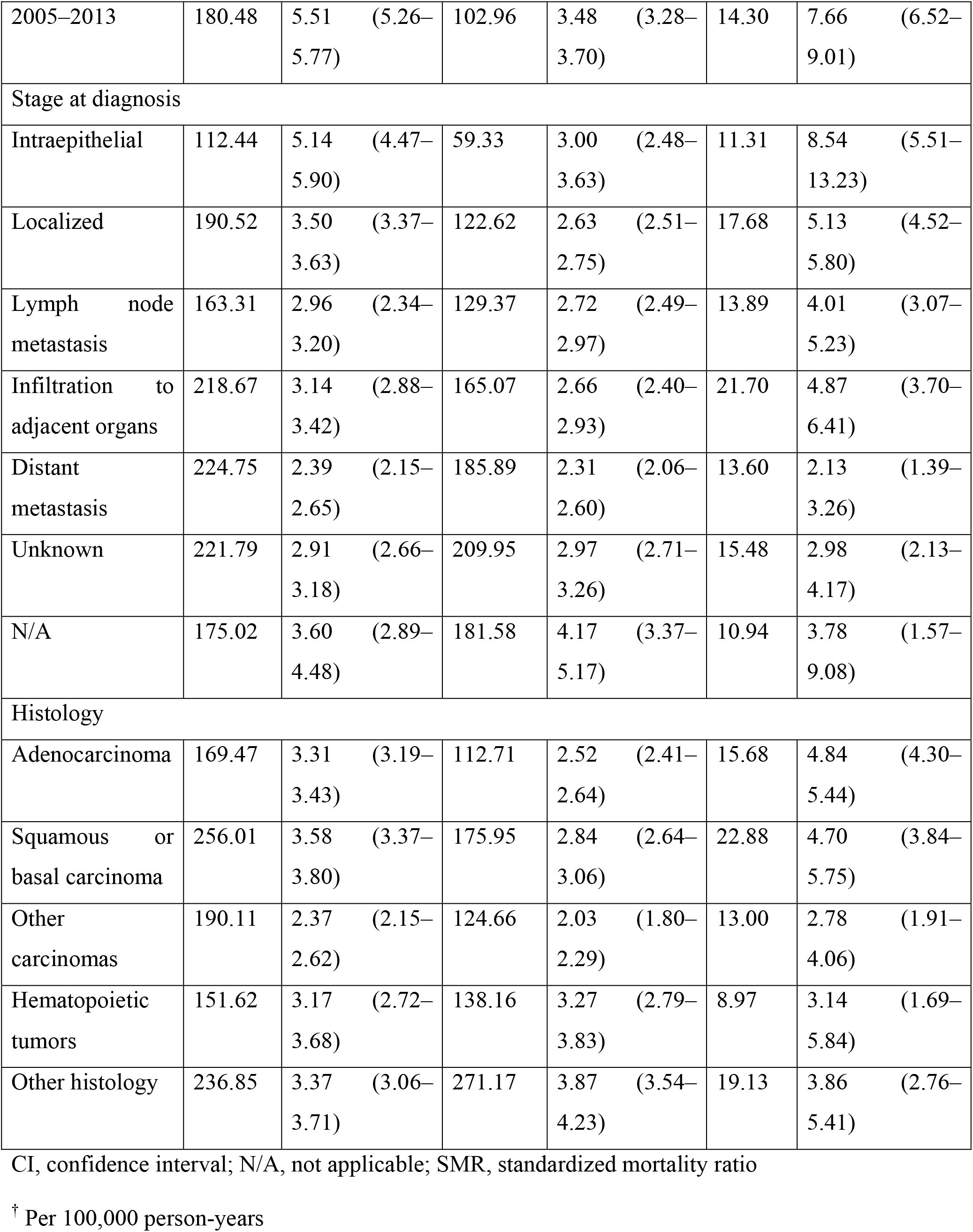
Standardized mortality ratios for ischemic heart disease, heart failure, and hypertensive disease in patients with cancer compared to the general population.

### SMR for heart disease according to cancer site

Figure 1. shows the SMRs for heart disease according to cancer site. The details are presented in **Supplemental Table S5**. The SMR was the highest in brain cancer, followed by that in bone, esophageal, uterine, and skin cancers. Meanwhile, the SMR was the lowest in liver cancer, followed by that in gallbladder, pancreatic, lung, and ovarian cancers.

**Figure 1.**
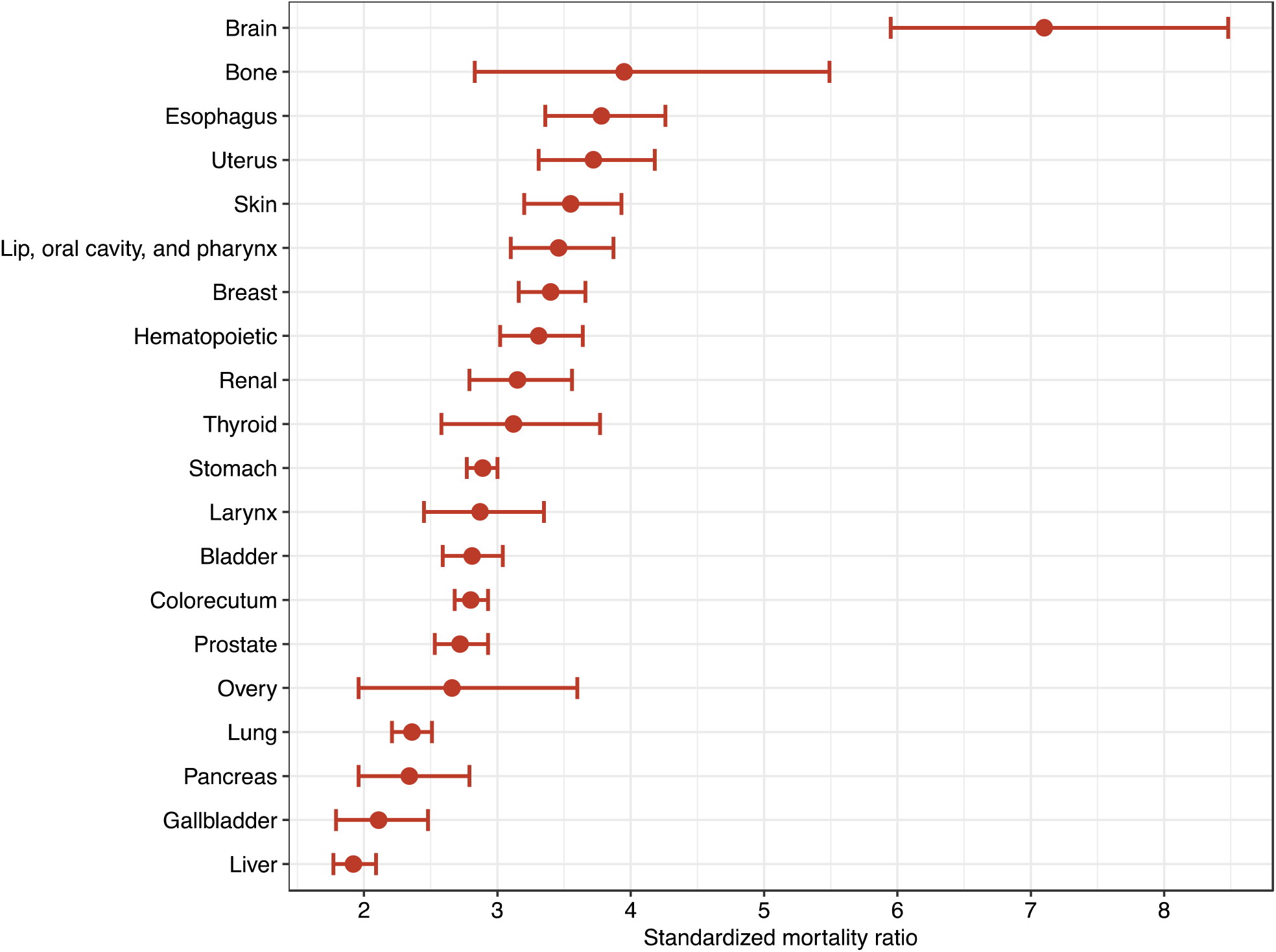
Standardized mortality ratio for heart disease death stratified by cancer site. The ordinate shows the cancer site, and the abscissa indicates the standardized mortality ratio (SMR) for heart disease-related death. Error bars show the 95% confidence interval. The plots are in the order of increasing SMR. Patients with cancer have a higher SMR for heart disease-related death than the general population.

### Risk of heart disease death according to time after cancer diagnosis

Figure 2. shows the SMR trend after cancer diagnosis. The details are presented in **Supplemental Tables S6–S9**. The SMR for heart disease was 3.28 (95% CI, 3.10–3.47) within the first 3 months after cancer diagnosis, declined once, and then gradually increased thereafter. The trend of SMR for ischemic heart disease and heart failure was similar to that of the SMR for overall heart disease. The SMRs for hypertensive disease varied with time after cancer diagnosis; however, in general, the SMRs increased over time. The SMR for hypertensive disease was the lowest within the first year (1.86 [95% CI, 1.19–2.92) and highest in the ninth year (9.63 [95% CI, 6.95–13.36]) after cancer diagnosis.

**Figure 2.**
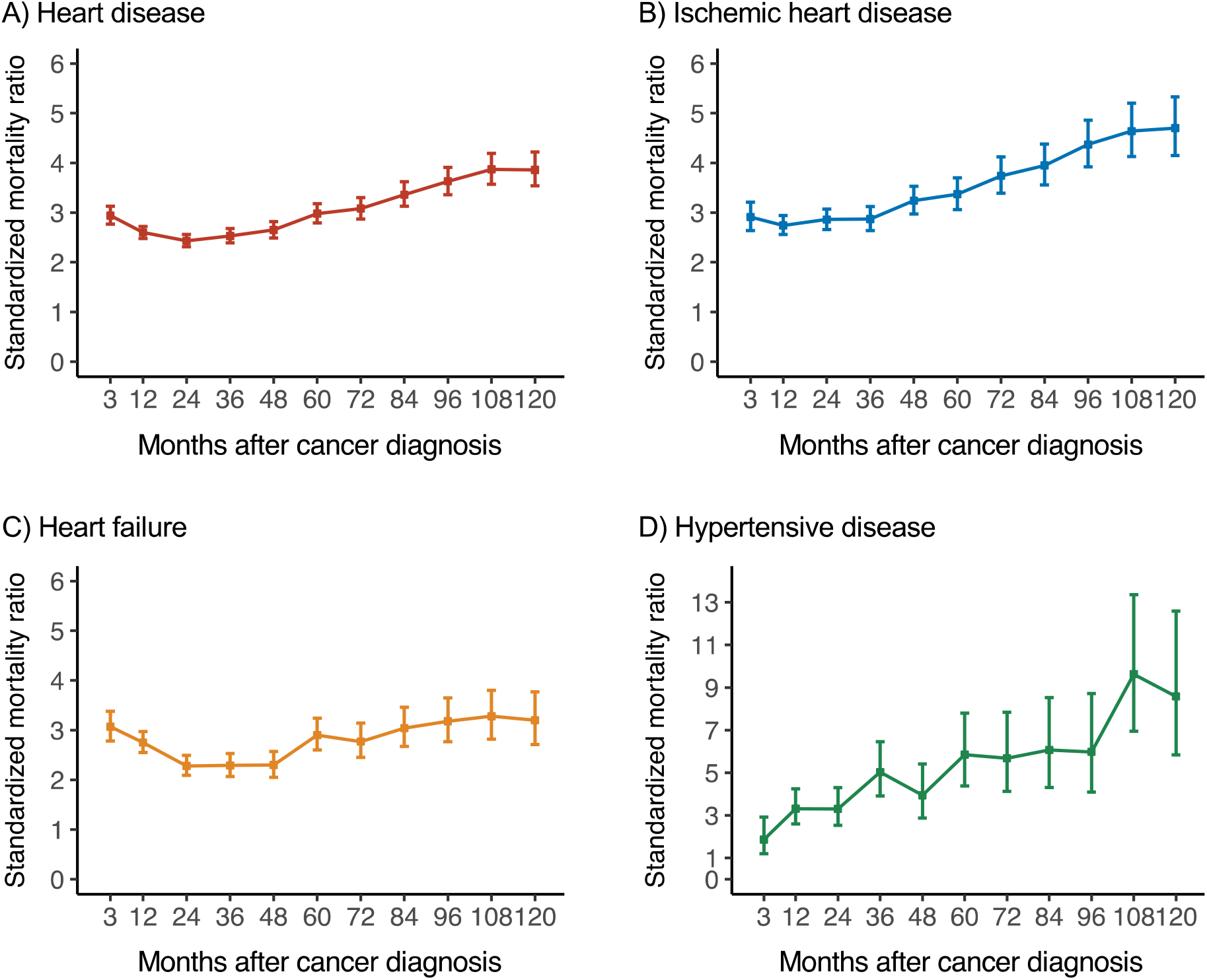
Trends in standard mortality ratio according to time after cancer diagnosis. The ordinate shows the standardized mortality ratio (SMR), and the abscissa indicates the time after cancer diagnosis. Error bars show the 95% confidence interval. (A) The SMR for heart disease tends to be high immediately after diagnosis, declines once, and then increases with time. (B, C) The SMR for ischemic heart disease and heart failure shows a trend similar to that of heart disease. (D) In the analysis of hypertensive disease, SMR varies with time, but they are generally increased over time.

### Risk of fatal heart disease by subgroup

**Table 3** shows the RRs of patients with cancer for fatal heart disease stratified by subgroup. Males were 1.4 times more likely to die from heart disease than females. RRs also gradually increased as the age at diagnosis increased. Meanwhile, RRs gradually decreased as the period of diagnosis became more recent. Patients with distant metastases had the highest RR among all cancer stages. **Table 4** shows the RRs for ischemic heart disease, heart failure, and hypertension. Males were more likely to die from ischemic heart disease, heart failure, and hypertensive disease than were females. RRs were also higher for patients who were older at diagnosis, while they were lower for those more recently diagnosed with ischemic heart disease, heart failure, and hypertensive disease. When analyzed according to stage at diagnosis, the RR for death from ischemic heart disease and heart failure was the highest for patients with distant metastasis. Meanwhile, for death from hypertensive disease, the RR was the highest for patients with cancer infiltration to adjacent organs. The RRs varied according to histological stage and heart disease.

**Table 3.** Relative risk of heart disease death in patients with cancer.

|  | RR | 95% CI | *P*-value |
| --- | --- | --- | --- |
| Sex |  |  |  |
| Female | 1.00 | – |  |
| Male | 1.39 | 1.34–1.45 | <0.001 |
| Age at diagnosis |  |  |  |
| ≤39 | 1.00 | – |  |
| 40–49 | 2.19 | 1.51–3.18 | <0.001 |
| 50–59 | 4.46 | 3.19–6.24 | <0.001 |
| 60–69 | 10.12 | 7.30–14.04 | <0.001 |
| 70–79 | 25.42 | 18.36–35.18 | <0.001 |
| ≥80 | 79.63 | 57.56–110.16 | <0.001 |
| Period of diagnosis |  |  |  |
| 1985–1994 | 1.00 | – |  |
| 1995–2004 | 0.60 | 0.57–0.63 | <0.001 |
| 2005–2013 | 0.51 | 0.48–0.53 | <0.001 |
| Stage at diagnosis |  |  |  |
| Intraepithelial | 1.00 | – |  |
| Localized | 1.29 | 1.17–1.43 | <0.001 |
| Lymph node metastasis | 1.46 | 1.31–1.63 | <0.001 |
| Infiltration to adjacent organs | 1.63 | 1.46–1.82 | <0.001 |
| Distant metastasis | 1.83 | 1.62–2.06 | <0.001 |
| Unknown | 1.64 | 1.47–1.84 | <0.001 |
| N/A | 2.82 | 2.30–3.46 | <0.001 |
| Histology |  |  |  |
| Adenocarcinoma | 1.00 | – |  |
| Squamous or basal carcinoma | 1.36 | 1.30–1.43 | <0.001 |
| Other carcinomas | 1.12 | 1.05–1.21 | 0.001 |
| Hematopoietic tumors | 0.99 | 0.87–1.13 | 0.94 |
| Other histology | 1.73 | 1.63–1.85 | <0.001 |

CI, confidence interval; N/A, not applicable; RR, relative risk

**Table 4.** Relative risk of ischemic heart disease, heart failure, and hypertensive disease death in patients with cancer.

|  | Ischemic heart disease |  |  | Heart failure |  |  | Hypertensive disease |  |  |
| --- | --- | --- | --- | --- | --- | --- | --- | --- | --- |
|  | RR | 95%<br>CI | *P*-<br>value | RR | 95%<br>CI | *P*-value | RR | 95%<br>CI | *P*-<br>value |
| Sex |  |  |  |  |  |  |  |  |  |
| Female | 1.00 | – |  | 1.00 | – |  | 1.00 | – |  |
| Male | 1.71 | 1.61–<br>1.82 | <0.001 | 1.19 | 1.12–<br>1.28 | <0.001 | 1.20 | 0.99–<br>1.45 | 0.061 |
| Age at diagnosis |  |  |  |  |  |  |  |  |  |
| ≤39 | 1.00 | – |  | 1.00 | – |  | – | – |  |
| 40–49 | 2.71 | 1.30–<br>5.65 | 0.008 | 1.72 | 1.00–<br>2.96 | 0.051 | – | – |  |
| 50–59 | 6.87 | 3.51–<br>13.42 | <0.001 | 3.04 | 1.88–<br>4.91 | <0.001 | 1.00 | – |  |
| 60–69 | 17.06 | 8.83–<br>32.97 | <0.001 | 6.14 | 3.87–<br>9.75 | <0.001 | 6.38 | 2.54–<br>16.01 | <0.001 |
| 70–79 | 42.37 | 21.97–<br>81.70 | <0.001 | 16.5<br>2 | 10.46<br>–<br>26.08 | <0.001 | 20.0<br>1 | 8.20–<br>48.86 | <0.001 |
| ≥80 | 109.2<br>1 | 56.65–<br>210.52 | <0.001 | 68.0<br>0 | 43.16<br>–<br>107.1<br>3 | <0.001 | 74.8<br>6 | 30.83<br>–<br>181.7<br>3 | <0.001 |
| Period of diagnosis |  |  |  |  |  |  |  |  |  |
| 1985–1994 | 1.00 | – |  | 1.00 | – |  | 1.00 | – |  |
| 1995–2004 | 0.78 | 0.73–<br>0.84 | <0.001 | 0.32 | 0.30–<br>0.35 | <0.001 | 0.64 | 0.50–<br>0.80 | <0.001 |
| 2005–2013 | 0.64 | 0.60–<br>0.69 | <0.001 | 0.29 | 0.27–<br>0.31 | <0.001 | 0.47 | 0.37–<br>0.60 | <0.001 |

| Stage at diagnosis |  |  |  |  |  |  |  |  |  |
| --- | --- | --- | --- | --- | --- | --- | --- | --- | --- |
| Intraepithelial | 1.00 | – |  | 1.00 | – |  | 1.00 | – |  |
| Localized | 1.30 | 1.12–<br>1.51 | <0.001 | 1.40 | 1.15–<br>1.71 | 0.001 | 1.14 | 0.72–<br>1.81 | 0.58 |
| Lymph node metastasis | 1.39 | 1.18–<br>1.64 | <0.001 | 1.76 | 1.42–<br>2.18 | <0.001 | 1.09 | 0.65–<br>1.84 | 0.74 |
| Infiltration to adjacent organs | 1.50 | 1.27–<br>1.77 | <0.001 | 1.94 | 1.56–<br>2.42 | <0.001 | 1.45 | 0.86–<br>2.44 | 0.16 |
| Distant metastasis | 1.63 | 1.36–<br>1.94 | <0.001 | 2.33 | 1.85–<br>2.93 | <0.001 | 1.01 | 0.54–<br>1.88 | 0.98 |
| Unknown | 1.45 | 1.22–<br>1.72 | <0.001 | 2.02 | 1.62–<br>2.51 | <0.001 | 0.97 | 0.55–<br>1.72 | 0.92 |
| N/A | 2.34 | 1.68–<br>3.26 | <0.001 | 3.96 | 2.74–<br>5.71 | <0.001 | 1.95 | 0.54–<br>7.08 | 0.31 |
| Histology |  |  |  |  |  |  |  |  |  |
| Adenocarcinoma | 1.00 | – |  | 1.00 | – |  | 1.00 | – |  |
| Squamous or basal carcinoma | 1.40 | 1.31–<br>1.51 | <0.001 | 1.43 | 1.31–<br>1.56 | <0.001 | 1.31 | 1.03–<br>1.66 | 0.03 |
| Other carcinomas | 1.11 | 1.00–<br>1.23 | 0.059 | 1.22 | 1.07–<br>1.39 | 0.003 | 0.89 | 0.60–<br>1.32 | 0.56 |
| Hematopoietic tumors | 0.91 | 0.74–<br>1.12 | 0.37 | 1.02 | 0.81–<br>1.29 | 0.88 | 0.65 | 0.27–<br>1.55 | 0.33 |
| Other histology | 1.56 | 1.40–<br>1.74 | <0.001 | 2.17 | 1.95–<br>2.41 | <0.001 | 1.37 | 0.95–<br>1.99 | 0.09 |

CI, confidence interval; N/A, not applicable; RR, relative risk

## Discussion

The risk of heart disease-related mortality in patients with cancer has not been elucidated to date. This study found the following findings. First, cancer survivors have a 2.9 times greater risk of heart disease death than the general population. Second, this risk varied by heart disease type: 3.3 times higher for ischemic heart disease, 2.7 times higher for heart failure, and 4.5 times higher for hypertensive disease. Third, the risk of death from heart disease tended to increase with time after cancer diagnosis. Finally, the risk of fatal heart disease among cancer survivors has decreased in recent years.

There have been studies on the risk of heart disease-related mortality among cancer survivors^6–14^. We found similar associations in the current analysis: patients with cancer had a 2.9 times higher risk of death from heart disease than the general population. The results were similar in the subgroup analysis by sex, age at diagnosis, year of diagnosis, and stage at diagnosis. However, some results were in contrast to previous findings. For example, the SMR of lung cancer was not as high as that in a previous study^12^. Additionally, while the SMR by stage at diagnosis was the highest in distant metastases and lowest in localized cancers in previous research, opposite findings were noted in our analysis. The SMR was the lowest in distant metastasis and highest in intraepithelial cancer. However, this result was similar to that of our previous study on stroke deaths among cancer survivors^18^. Although the SMRs could not be compared due to the different populations studied, these findings are noteworthy in the context of prognosis and management of heart disease risk in cancer survivors. The factors that influenced these results are unclear, but regional differences and the extent of malignancies are possible contributing factors.

Several mechanisms explain the relationship between cancer and heart disease. Lifestyle factors such as diabetes and smoking are associated with the development of both cancer and heart disease^4,5,22^. Neuroendocrine factors, oxidative stress, inflammatory cytokines, and impaired immune system have been implicated in the increased risk of developing heart disease in cancer patients ^4,5^. Cancer-related coagulation abnormalities can cause arterial thromboembolisms, including myocardial infarction^23^. Certain anticancer agents, including anthracyclines^24^, tyrosine kinase inhibitors^25^, and immune checkpoint inhibitors^26,27^, have cardiotoxic adverse effects. Further, late-onset toxicity has been found to be associated with an increased risk of heart disease-related mortality among cancer survivors^28^. An increased risk of cardiovascular events with surgery^29^ and radiotherapy^30^ has also been reported.

In this study, the risk of ischemic heart disease-related death was 3.3 times greater in patients with cancer than in the general population. Paterson et al. assessed the cardiovascular risk in more than 220,000 patients with cancer. The HR (95% CI) of death from myocardial infarction was 1.01 (0.97–1.05), which did not differ from that in the general population^31^. The strength of their study is that the median observation period was >10 years, and the analysis was more in depth, adjusting for vascular risk. Our study also analyzed the risk of death from ischemic heart disease using data from more than 500,000 patients with cancer, but it was limited by a median observation period of approximately 4 years and failure to adjust for the vascular risk factors such as hypertension and diabetes mellitus. Cancer patients have been established to have a high risk of ischemic heart disease^32–34^. Further studies are needed to determine whether cancer survivors have an elevated risk of mortality from ischemic heart disease.

The current study found that the risk of fatal heart disease increased over time after cancer diagnosis. Notably, the risk was immediately high after cancer diagnosis. This trend may be due to cancer treatment initiated after diagnosis. Surgery increases the risk of major adverse cardiovascular disease^29^, and a particular type of chemotherapy can cause vascular toxicity^24–27^. Cancer survivors develop a wide range of radiation-related cardiotoxic complications^30^. The subsequent increase in the risk of heart disease mortality over time may be due to a relative increase in cardiac mortality against cancer-related deaths. Collectively, these findings support that cancer treatment may be associated with an increase in the SMR after cancer diagnosis.

When examining the SMR by cancer site, the SMR from fatal heart disease was the highest for brain cancers. One possible explanation for the increased risk of death from heart disease in brain tumors is the brain-heart connection^35^. Damage to the brain parenchyma affects the balance of neuromodulating hormones, triggering arrhythmias and impairing cardiac function^35^. Brain tumors can cause fatal neurogenic cardiac diseases. Reports of an increased risk of cardiovascular mortality in patients with brain tumors would support this hypothesis^36^.

However, our analyses may have overestimated the risk of death from heart disease^37^. The causes of death in the NANDE database are collected through official Japanese statistics reports, which register the cause of death from death certificates. When a brain tumor patient dies suddenly, the cause of death may be listed as heart disease because the cause of death is unknown. There may also be instances in which the cause of death is listed as heart disease in patients with terminal disease.

The strength of this study is that it assessed the risk of heart disease mortality among cancer survivors using a large cancer registry that spanned approximately 30 years. However, this study also has some limitations. First, risks were analyzed without adjustment for confounding variables such as vascular risk factors. Second, advances in cancer treatment apparently influenced prognosis, but information regarding treatment was unavailable. Third, we excluded cases with incomplete information concerning survival days or diagnosis dates, death certificate notification, or death certificate only, as well as cases of simultaneous or synchronous cancers. Approximately 3.4% of patients were lost to follow-up. When comparing cancer types between patients who were included, excluded, and lost to follow-up, the proportions were similar for many cancer types. However, colorectal, breast, uterine, and prostate cancers were more common in the study population. In contrast, liver, gallbladder, pancreatic, and lung cancers were more common in the excluded population and those lost to follow-up. This may have resulted in selection bias. Finally, a follow-up period of 10 years after cancer diagnosis may be inadequate to investigate heart disease-related deaths.

In conclusion, cancer survivors have a high risk of heart disease-related mortality. Thus, clinicians should recognize the risk of fatal heart disease in cancer patients. Careful clinical management, including of both cancer and heart disease, will improve quality of life and thus be beneficial to cancer survivors.

## Data Availability

Data are available on reasonable request to the corresponding author.

## Abbreviations

CIs: confidence intervals
ICD: International Classification of Diseases
NANDE: Neoplasms ANd other causes of DEath
OCR: Osaka Cancer Registry
RR: relative risk
SEER: Surveillance, Epidemiology, and End Results
SMRs: standardized mortality ratios

## Acknowledgments

None

## Data availability

Data are available on reasonable request to the corresponding author.

## Ethics approval and consent to participate

Informed consent was waived owing to the retrospective nature of the study. The Institutional Review Board of Osaka University, Suita, Japan, approved the study protocol (approval number: 17315-3).

## Author Contributions

Y.G., H.M., T. Sobue and I.M. contributed to the conception and design of the study. Y.G., L.Z., T.M., Y.O., T. Sobue, and I.M. contributed to the data acquisition and analysis. Y.G. and L.Z. contributed to drafting a significant portion of the manuscript or figures. L.Z., T.M., T. Sasaki, Y.O., H.M., T. Sobue, and I.M. revised the manuscript critically for important intellectual content. All the authors approved the final manuscript.

